# Computer Vision Analysis of Specimen Mammography to Predict Margin Status

**DOI:** 10.1101/2023.03.06.23286864

**Authors:** Kevin A Chen, Kathryn E Kirchoff, Logan R Butler, Alexa D Holloway, Muneera R Kapadia, Kristalyn K Gallagher, Shawn M Gomez

**Author notes:** Correspondence should be addressed to SMG and KKG.

## Abstract

Intra-operative specimen mammography is a valuable tool in breast cancer surgery, providing immediate assessment of margins for a resected tumor. However, the accuracy of specimen mammography in detecting microscopic margin positivity is low. We sought to develop a deep learning-based model to predict the pathologic margin status of resected breast tumors using specimen mammography. A dataset of specimen mammography images matched with pathology reports describing margin status was collected. Models pre-trained on radiologic images were developed and compared with models pre-trained on non-medical images. Model performance was assessed using sensitivity, specificity, and area under the receiver operating characteristic curve (AUROC). The dataset included 821 images and 53% had positive margins. For three out of four model architectures tested, models pre-trained on radiologic images outperformed domain-agnostic models. The highest performing model, InceptionV3, showed a sensitivity of 84%, a specificity of 42%, and AUROC of 0.71. These results compare favorably with the published literature on surgeon and radiologist interpretation of specimen mammography. With further development, these models could assist clinicians with identifying positive margins intra-operatively and decrease the rate of positive margins and re-operation in breast-conserving surgery.

## Introduction

Breast-conserving surgery is the standard of care for early-stage breast cancer, balancing oncologic resection and cosmesis.^1^ For breast-conserving surgery, obtaining negative margins is critical in reducing local recurrence, and reported re-operation rates for positive margins remain high, at about 22%.^2,3^ Margin status is determined post-operatively with surgical pathology, with results days to weeks after completion of the surgery. Multiple interventions have attempted to identify positive margins intra-operatively, including specimen mammography, which involves taking an X-ray of the resected tumor using a portable scanner in the operating room.

Specimen mammography is a widely used technique that provides immediate feedback on the quality of resection and may assist surgeons with identifying suspicious margins and directing targeted removal of additional tissue.^4^ However, specimen mammography can be inaccurate, with sensitivity ranging from 20% to 58% and area under the receiver operating characteristic curve (AUROC) ∼0.7.^5–8^ A tool to assist clinicians with interpretation of specimen mammography for partial mastectomy would be helpful to assist identification of positive margins and reduce the rate of re-operation. This tool would be particularly helpful for low-volume and low-resource centers which see higher rates of positive margins and may have less familiarity with interpreting specimen mammography.^9^

Deep learning has been extensively applied to screening mammography, with a recent systematic review identifying 82 relevant studies.^10^ Deep learning also shows promise for intra-operative use, with recent applications to the interpretation of laparoscopic video.^11–13^ However, deep learning has not yet been applied to intra-operative specimen mammography.

The goal of this study is to develop a model that can accurately predict margin positivity for partial mastectomy based on specimen mammography alone. We sought to use pre-training specific to radiology through the RadImageNet project to improve transfer learning.^14^ We hypothesized that pre-training with a medical image dataset would result in a more accurate model that could match previously published metrics for human accuracy in predicting margin status.

## Methods

### Study design and outcomes

Prior to data collection, approval from the University of North Carolina Institutional Review Board (#20-1820) was obtained. Written informed consent was not required because data was collected retrospectively and risks to patient privacy were minimized. Data was collected in two phases. First, from 7/2017 to 6/2020, specimen mammograms were prospectively collected as part of a single surgeon’s quality assurance process. Second, from 8/2020 to 4/2022, specimen mammograms from four surgeons were collected specifically for this project. Using retrospective chart review, specimen mammograms were matched with pathology reports and categorized into positive and negative classes based on NCCN guidelines.^15^ For specimens with invasive cancer, a positive margin was defined as “ink on tumor.” For specimens with DCIS, a positive margin was defined as DCIS within 2mm of the margin. For specimens containing both DCIS and invasive cancer, but with DCIS within 2mm of the margin, we elected to categorize this pathology as positive, while acknowledging that, clinically, this result is treated as negative. This is because margin positive DCIS in this situation is unlikely to appear different compared with DCIS alone. In addition, margin assessment was performed on the original specimen, excluding cavity shave margins, as these are not imaged by specimen mammography. In the second phase of data collection, additional clinical information was collected including age, race/ethnicity, BMI, breast density, tumor type, tumor grade, and tumor size.

### Data processing

The dataset was divided randomly into training, validation, and test sets in a 60/20/20 ratio. A single anterior-posterior image was selected for each specimen and resized to 512 × 512 pixels. The training set underwent standard data augmentation including random flipping (vertically and horizontally), zoom, shifts (horizontal and vertical), and rotation.^16^

### Modeling

We developed models based on architectures used by the RadImageNet project, including ResNet-50, InceptionV3, Inception-ResNet-v2, and DenseNet-121. The RadImageNet models are pretrained on 1.35 million annotated medical images.^14^ For comparison, we developed models using the same architectures, pre-trained on ImageNet, which largely consists of non-medical images. For each model, two additional fully connected layers, followed by dropout layers, and an output classification layer were added. A diagram of the model is shown in Figure 1. The number of fully connected layers, neurons, and dropout were held constant to facilitate comparison. After the highest performing model and pre-training dataset were identified, the number of fully connected layers, neurons, dropout, and learning rate were tuned on the training/validation sets.

**Figure 1.**
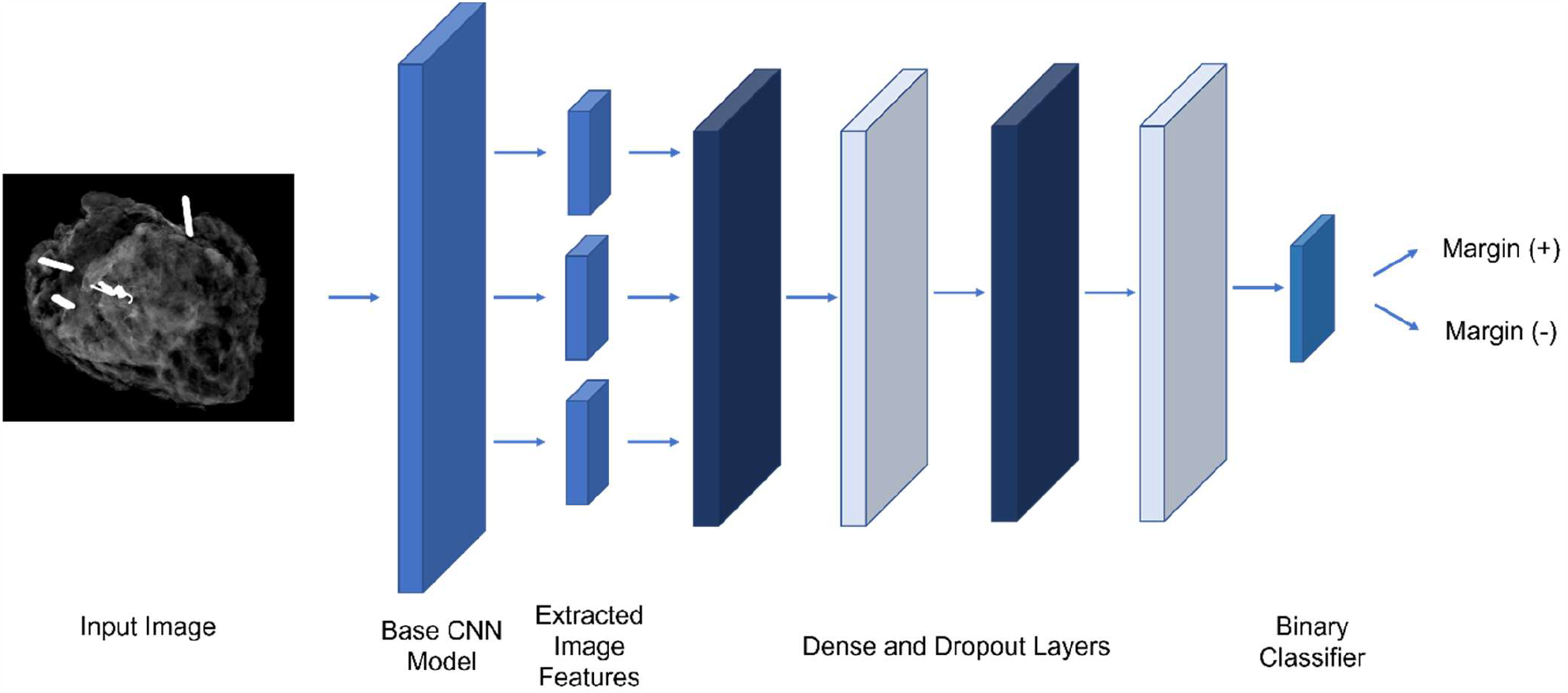
Diagram of model structure

For all comparisons, transfer learning was completed in two phases. First, the base architecture was frozen and the model was trained at a higher learning rate. Second, the base architecture was unfrozen and trained at a lower learning rate. Early stopping was used to avoid overfitting.^17^

### Model Evaluation

Model performance was assessed using sensitivity, specificity, positive predictive value (PPV), negative predictive value (NPV), area under the receiver operating characteristic curve (AUROC), and area under the precision-recall curve (AUPRC). For the highest performing model (based on AUROC), evaluation metrics were calculated for categories within breast density and race/ethnicity because screening mammography has previously been shown to have different accuracy within these groups.^18,19^ In addition, Grad-CAM was used to create saliency maps to assess pixel importance for 10 randomly selected images from the test set and these were visually assessed.^20^ Figure 2 shows the overall process for model development and evaluation.

**Figure 2.**
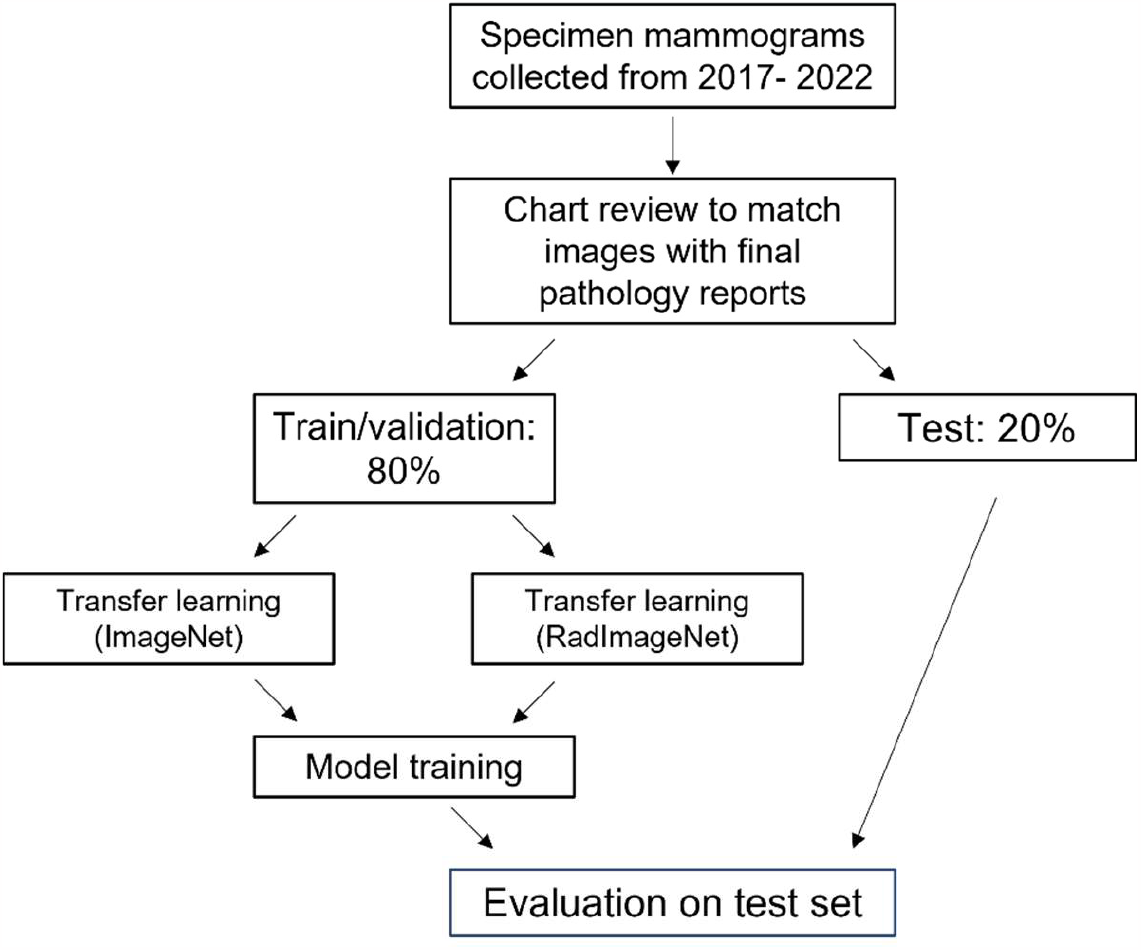
Flowchart of study design

Models were implemented using Python (version 3.8) and the scikit-learn and Tensor-flow/Keras libraries.^21,22^ For characterization of the cohort, Chi-squared test was used to compare categorical variables and T-test to compare continuous variables. Code to reproduce this work is available at github.com/gomezlab/cvsm.

## Results

### Study population characteristics

A The dataset included 821 images. Of these, 450 were collected in the first, single surgeon phase, while 371 were collected in the second, multiple surgeon phase. 431 (52.5%) images had positive margins. Representative images for each classification are shown in Figure 3. Within the positive classification, 128 had mixed pathology with both invasive cancer and DCIS 2mm from the margin. The average age was 60 and most patients had IDC (67.1%) or DCIS (20.5%). Non-Hispanic White patients comprised 70.1% of the cohort, compared with 18.1% Non-Hispanic Black and 7.8% Hispanic. Most patients had a breast density of B (45.8%) or C (36.9%). Patients with positive margins were slightly older and more likely to have a tumor grade of 2 (Table 1). After splitting into training, validation, a nd testing groups, 492 images were used for training, 165 for validation, and 164 for testing.

**Table 1.** Demographic and clinical characteristics of patients included in the dataset

|  |  | Margin (-) | Margin (+) | P-Value |
| --- | --- | --- | --- | --- |
| **n** |  | 148 | 223 |  |
| **Age, mean (SD)** |  | 59.2 (12.8) | 61.6 (13.2) | 0.081 |
| **Race/Ethnicity, n (%)** | **Asian** | 7 (4.7) | 5 (2.2) | 0.118 |
|  | **Hispanic** | 9 (6.1) | 20 (9.0) |  |
|  | **Non-Hispanic Black** | 25 (16.9) | 42 (18.8) |  |
|  | **Non-Hispanic White** | 104 (70.3) | 156 (70.0) |  |
|  | **Other/Unknown** | 3 (2.0) |  |  |
| **BMI, mean (SD)** |  | 28.9 (6.7) | 28.9 (6.4) | 1.000 |
| **Density, n (%)** | **A** | 6 (4.1) | 11 (4.9) | 0.573 |
|  | **B** | 63 (42.6) | 107 (48.0) |  |
|  | **C** | 61 (41.2) | 76 (34.1) |  |
|  | **D** | 18 (12.2) | 29 (13.0) |  |
| **Tumor Grade, n (%)** | **0.0** | 11 (7.4) | 18 (8.1) | 0.034 |
|  | **1.0** | 34 (23.0) | 50 (22.4) |  |
|  | **2.0** | 51 (34.5) | 105 (47.1) |  |
|  | **3.0** | 52 (35.1) | 50 (22.4) |  |
| **Tumor Size, mean (SD)** |  | 7.4 (3.3) | 7.6 (3.1) | 0.610 |
| **Tumor Type, n (%)** | **DCIS** | 30 (20.3) | 46 (20.6) | 0.588 |
|  | **EPC** | 4 (2.7) | 3 (1.3) |  |
|  | **IDC** | 96 (64.9) | 153 (68.6) |  |
|  | **ILC** | 13 (8.8) | 18 (8.1) |  |
|  | **LCIS** | 5 (3.4) | 3 (1.3) |  |

DCIS – Ductal carcinoma in situ, EPC – Encapsulated papillary carcinoma, IDC – Invasive ductal carcinoma, ILC – invasive lobular carcinoma, LCIS – lobular carcinoma in situ

**Figure 3.**
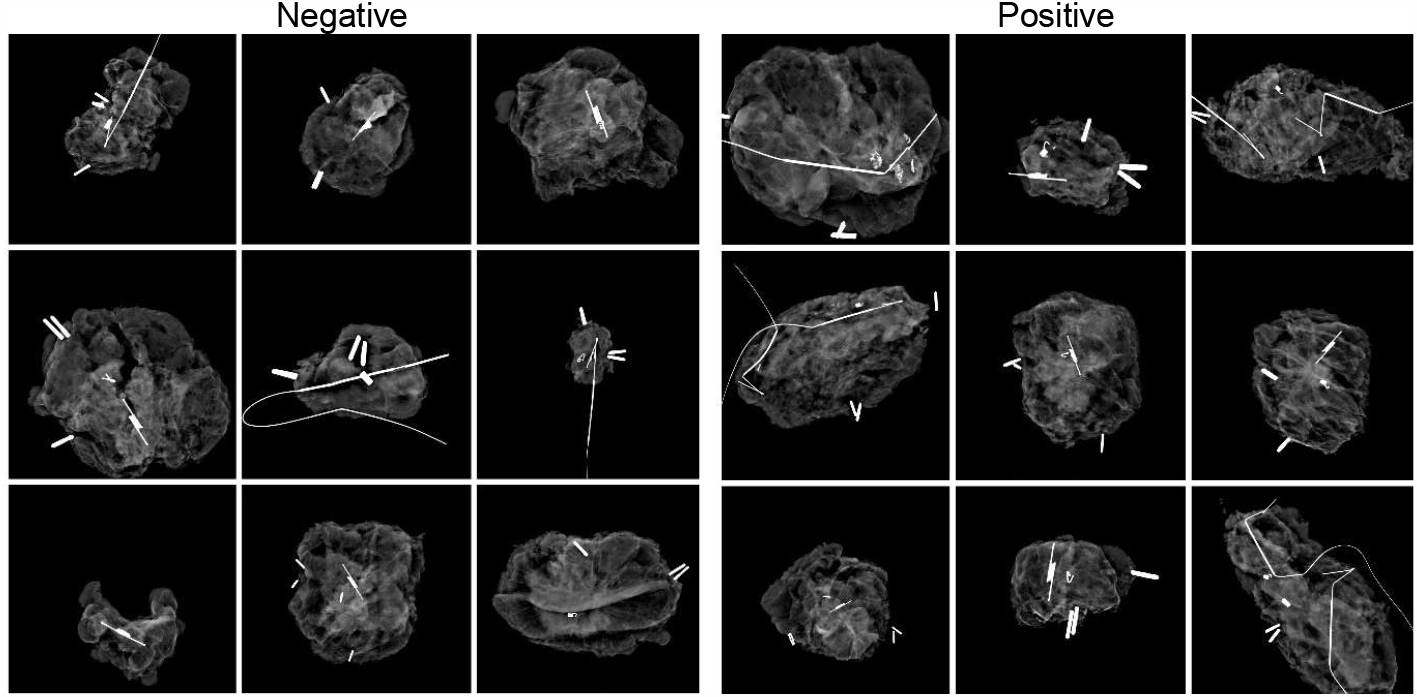
Example specimen mammograms by margin status class

We first compared the performance of models pre-trained on RadImageNet, compared with models pre-trained on ImageNet. Overall, RadImageNet models showed higher accuracy compared with ImageNet models with AUROC 0.63 - 0.71 vs. 0.46 - 0.68, respectively. All architectures showed higher performance with RadImageNet pre-training except for DenseNet121. For this model, performance between the two pre-training regimens was similar at 0.66 for RadImageNet compared with 0.68 for ImageNet. The highest performing model was InceptionV3 with RadImageNet pre-training. Receiver operating characteristic curves and precision-recall curves are shown for all models in Figure 4. Based on this analysis, we performed hyperparameter tuning on InceptionV3 with RadImageNet pre-training. After hyperparameter tuning, this model showed an AUROC of 0.72, AUPRC of 0.70, sensitivity of 84%, specificity of 42%, positive predictive value of 62%, and negative predictive value of 70%. In subset analysis, we found that model performance was worse for patients with dense breast tissue (Category D) compared with less dense tissue (Categories A-C) (Table 2). For race/ethnicity, model performance was worse for White Non-Hispanic patients compared with non-White patients, although this appeared to be partly driven by breast density, with 14% of Non-Hispanic White patients having Category D density, compared with 9% of non-White patients (Table 3).

**Table 2.**
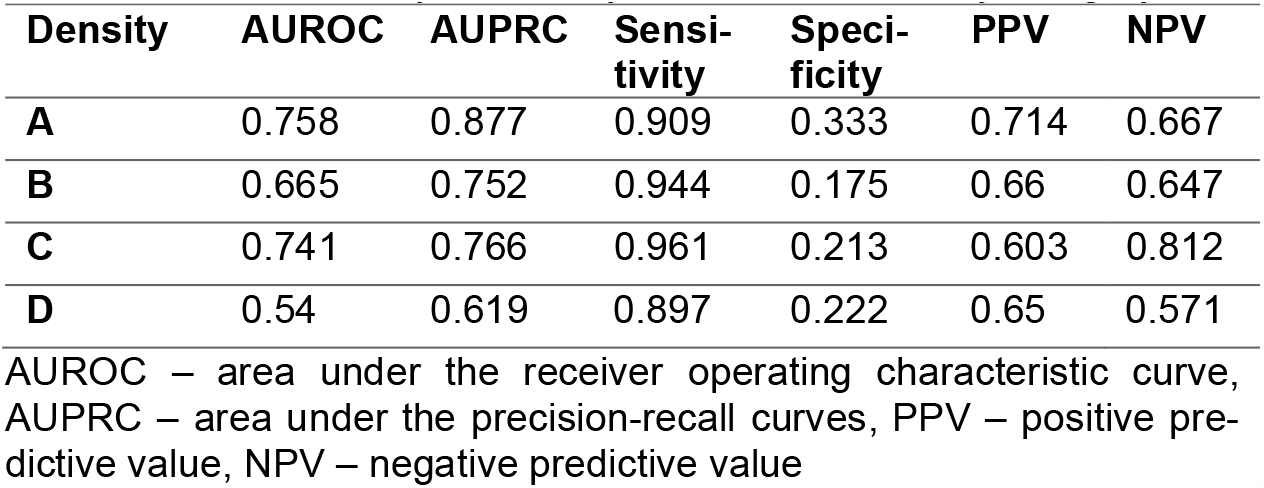
Model accuracy metrics by breast tissue density category

**Figure 4.**
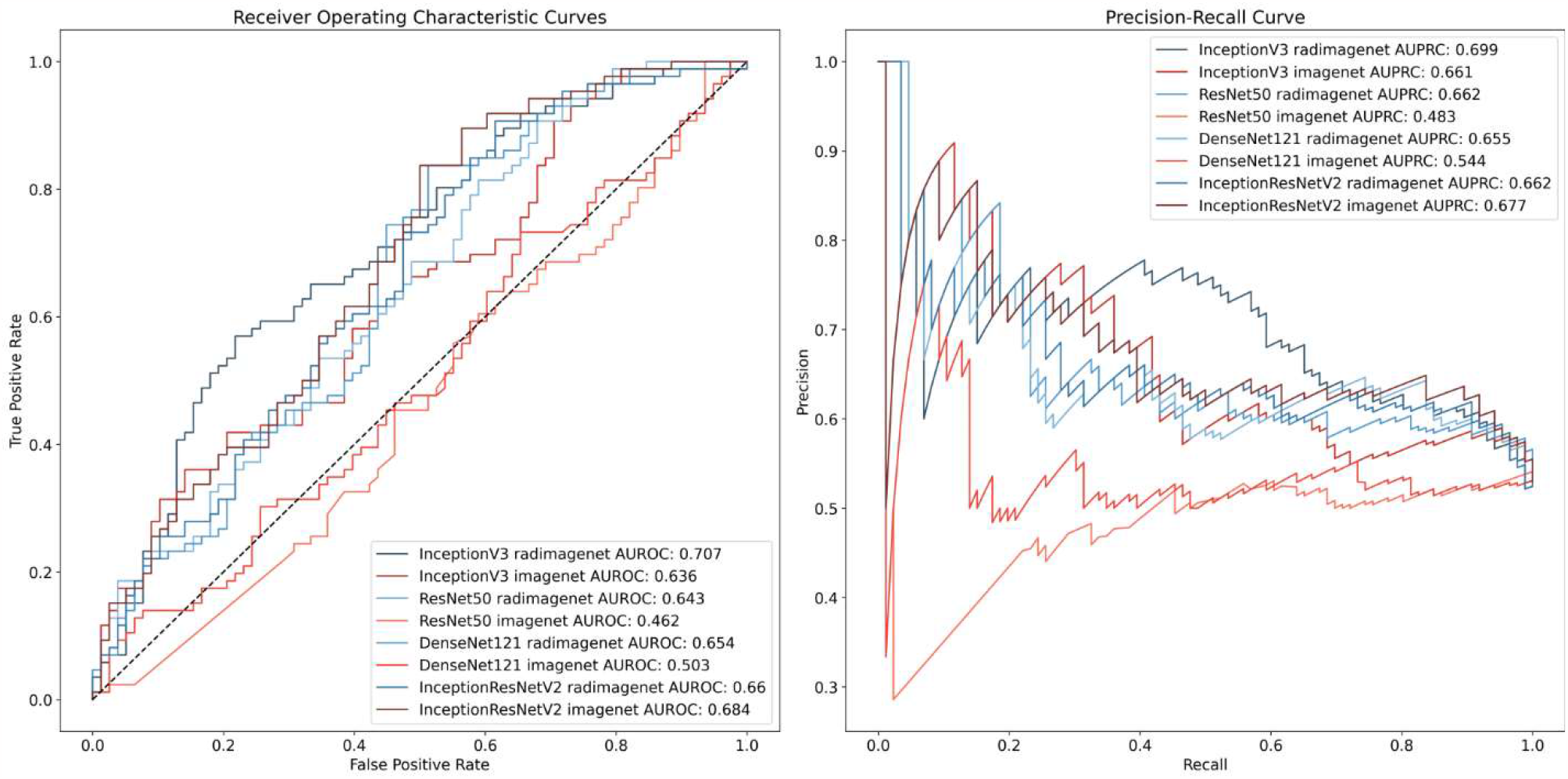
Receiver operating characteristic and precision-recall curves for models predicting margin status

To better understand what image features contributed the most to model decision-making, we assessed where the attention of the model was focused. Pixel importance analysis using Grad-CAM showed that, in most cases, the InceptionV3 model trained on specimen mammography images had attention focused on relevant parts of the image, such as localization wires, biopsy clips, and areas of tumor, while the InceptionV3 model trained on natural images did not (Figure 5).

**Figure 5.**
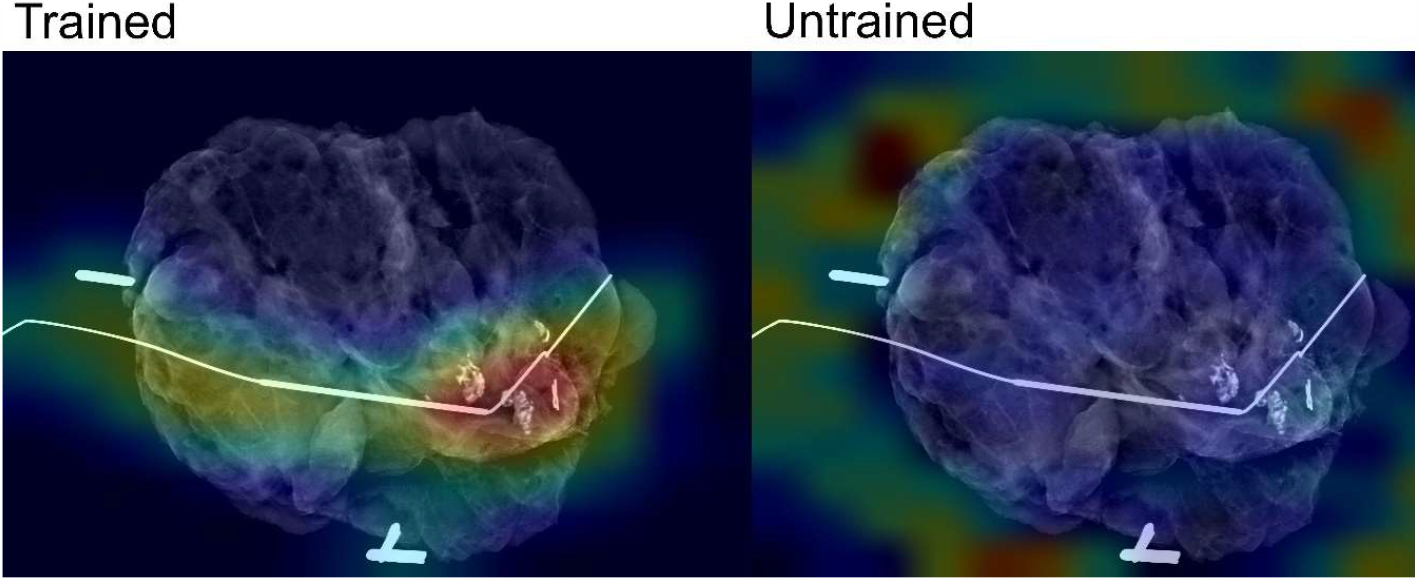
Analysis of pixel importance using Grad-CAM for trained and untrained InceptionV3 models

## Discussion

This project developed a prototype deep learning model which assesses the margin status of specimen mammograms. Pre-training with radiology images was found to improve model predictions compared with natural images. With an internal test set, model accuracy compared favorably with human interpretation, with a sensitivity of 84%, a specificity of 42%, and AUROC of 0.71.

Despite advances in intra-operative margin assessment, the rate of positive margins after partial mastectomy remains high.^3^ Specimen mammography is a widely used method that benefits from its availability within the operating room and its ability to provide immediate feedback. However, the diagnostic accuracy of specimen mammography is highly variable and relatively low, with reported sensitivity ranging from 20-58%.^5–8^ A 2017 meta-analysis of 9 studies showed a pooled sensitivity of 53% (95% CI 45 – 61%) with a pooled specificity of 84% (95% CI 77 - 89%).^23^ AUROC is similarly variable, but low, ranging from 0.60 to 0.73.^23–27^ The current models’ accuracy metrics are higher than many of the results published in the literature, demonstrating the potential of this approach.

Another interesting finding from our study was that our model showed lower accuracy in predicting margin status among patients with the highest breast density (Category D). This is expected, given similar findings for screening mammography, but has not been previously reported for specimen mammography.^19^ In contrast to previous studies applying artificial intelligence (AI) to mammography, our models show higher accuracy among non-White patients compared with White patients.^18^ This could be because non-White patients are well-represented within our dataset and because of the higher percentage of Category D breast density among White patients.

More generally, the overall performance of the current models mirror other recent advances in computer vision classification of mammograms, which show improved accuracy when radiologists are assisted by AI.^28–30^ AI has also recently been successfully applied to laparoscopic video, demonstrating its potential to assist with real-time, intra-operative decision-making.^31–33^ AI-assisted interpretation of specimen mammography may function similarly. It will be particularly useful for low-volume or low-resource centers which lack access to dedicated breast surgeons, radiologists, or pathologists and see higher rates of positive margins.^9^ AI-assisted clinical decision-support has the potential to improve equity in surgical outcomes for patients treated at these institutions.^34^ However, further development of the current models is needed, including collection of a larger, multi-institutional training set and mammogram-specific transfer learning.

This project has several limitations. The accuracy and generalizability of the model is most significantly limited by the size and single-institution nature of the dataset. A larger, multi-institutional dataset would likely result in a more accurate and robust model and verify its external validity. Second, the rate of positive margins is higher than previously reported.^3^ However, this is expected given our exclusion of cavity shave margins and focus on DCIS margins in mixed IDC/DCIS pathology. Third, the model does not identify which margin may be involved. Use of model attention techniques to automatically localize image features associated with a positive margin is possible and represents a target for future research.^35,36^ Newer imaging techniques, such as 3D tomosynthesis or optical coherence tomography may also be more accurate compared with specimen mammography and result in improved models.^37–39^ However, the current approach has the advantage of using widely available systems rather than new hardware, which may be costly. Finally, we did not assess radiologist or surgeon accuracy for margin classification on our dataset, although this is likely to be similar to previous literature.

## Conclusions

In conclusion, we developed a prototype model that can predict the pathologic margin status of partial mastectomy specimens based on intra-operative specimen mammography. The model’s predictions compare highly favorably with human interpretation. Optimized and externally validated versions of this model could assist surgeons with identifying positive margins intra-operatively and ultimately reduce the need for re-operation in breast-conserving surgery without requiring additional imaging hardware.

## Data Availability

All data produced in the present study are available upon reasonable request to the authors.

## Acknowledgements

This paper was typeset with the bioRxiv word template by @Chrelli: www.github.com/chrelli/bioRxiv-word-template

